# Wastewater metagenomic sequencing enables broad pathogen and resistome monitoring in Lagos, Nigeria

**DOI:** 10.64898/2026.09.22.26363641

**Authors:** Edyth Parker, Faith Oladipo, Joshua I. Levy, Oludayo O. Ope-Ewe, Harouna Soumare, Kristian Andersen, Anise N Happi, Christian T. Happi

## Abstract

Population-scale pathogen surveillance is limited worldwide, particularly in resource-constrained settings, where clinical systems only monitor select priority pathogens and reach only those who can access healthcare. Wastewater-based epidemiology addresses many of these gaps, offering cost-effective surveillance that is adaptable across diverse implementation contexts. In Lagos, Nigeria, a high enteric pathogen burden coincides with a complex sanitation landscape. However, circulating pathogen diversity in Lagos is largely uncharacterised outside of outbreak contexts. Here we characterize human-associated pathogen diversity in Lagos with virus-enriched metagenomic and metatranscriptomic sequencing of untreated wastewater sampled from open drainage canals from July-August 2024. We detected a diverse human-associated virome dominated by enteric adenoviruses, astroviruses and caliciviruses. We recovered numerous partial and near-complete genomes from pathogens of public health concern, including noroviruses, enteroviruses and sapoviruses. Although our enrichment targeted viruses, we detected diverse bacterial pathogens, including recurrent detection of *Vibrio cholerae*, other enteric bacteria, and zoonotic pathogens such as *Streptococcus suis.* We also detected antimicrobial resistance genes, including aminoglycoside, beta-lactam and fluoroquinolone resistance and clinically important genes such as *mcr*. Together, these results establish a regional baseline of pathogen diversity and demonstrate the utility of virus-enriched sequencing for integrated wastewater surveillance.

## Introduction

Recent outbreaks and pandemics have underscored the need for more comprehensive pathogen surveillance.^1,2^ To be effective, surveillance systems need to enable early detection, burden estimation, incidence monitoring and intervention prioritisation across diverse pathogens, while integrating with genomic epidemiology to characterise pathogen evolution and spread. Existing systems, however, are restricted to a small set of priority pathogens and rely on syndromic and clinical surveillance, which fails to capture asymptomatic infections and is biased by healthcare access.^3^ Wastewater-based epidemiology (WBE) addresses many of these limitations, providing cost-effective and non-invasive surveillance of the general population that can often detect pathogens earlier than clinical systems.^4,5^ WBE has successfully been applied to surveillance of endemic and emerging viruses, tracking of viral variants and monitoring of antimicrobial resistance genes, particularly for pathogens shed in stool, urine, respiratory secretions or other waste sources.^6–9^ WBE supports both targeted and untargeted approaches, such as PCR-based quantification of pathogen abundance and metagenomic sequencing.^10,11^ Targeted assays such as qPCR, ddPCR, and amplicon sequencing are highly sensitive for known pathogen detection, particularly at low concentrations in complex samples, but cannot detect novel, unexpected or divergent pathogens.^12,13^ Metagenomic or metatranscriptomic sequencing can characterize diverse viral, bacterial and fungal pathogens and resistance genes from a single sample, but sensitivity depends on pathogen titer and sequencing strategy.^12,14^ Although wastewater is a complex mixture of human-, non-human animal-and environmental-associated microbes, cross-sectional metagenomic sequencing of wastewater has the potential to characterize regional pathogen diversity and burden to help guide public health decision-making. However, wastewater surveillance, particularly using sequencing-based approaches, is restricted most of Africa, limiting understanding of local pathogen diversity and antibiotic resistance profiles.^6^

Lagos, Nigeria, is a global megacity of 16-21 million people, and a major trade, travel, and transport hub.^15^ In Lagos, rapid population growth and urbanisation have outpaced infrastructure development to produce a complex and fragmented sanitation landscape.^16–18^ Limited formal sewage treatment, septic systems and open canals converge across the city.^18^ These canals receive untreated domestic and commercial effluent, stormwater, and direct fecal contamination as they pass through densely populated communities, including input from domestic or peri-domestic animals and markets.^18^ The enteric pathogen burden is high, with canal overflow during heavy rainfall associated with seasonal diarrhoeal disease and cholera outbreaks.^18,19^ WBE is well suited to this context, as it can capture population-level data across socioeconomically diverse communities, including groups underrepresented in clinical surveillance.^20^ However, effective wastewater surveillance design for Lagos requires regional baselines for the wastewater virome, bacterial pathogens and AMR resistome, so targeted monitoring can focus on locally relevant organisms and genes.^6,7,9,10,21^

Here we establish a cross-sectional baseline of the human-associated pathogen diversity in untreated wastewater from a high-density urban setting in Lagos. Using virus-enriched metagenomic sequencing of untreated wastewater samples collected from the Lagos canal network, we recovered a diverse human-associated virome, including noroviruses and enteroviruses, bacterial pathogens including *Vibrio cholerae* and *Streptococcus suis* as well as antimicrobial resistance markers including aminoglycoside, beta-lactam and fluoroquinolone resistance. Our findings demonstrate the power of metagenomic and metatranscriptomic sequencing for integrated virome, bacteriome and resistome wastewater surveillance and its potential to improve our understanding of population-scale pathogen evolution and spread.

## Results

To detect and characterize pathogens of clinical concern in the wastewater of a high-density community in Lagos, we collected 38 grab samples of untreated wastewater from six sites in Oshodi/Isolo local government area (LGA) at three time points in late July to early August 2024 (Figure 1A, B). Oshodi/Isolo LGA is a densely populated, socioeconomically diverse area, with nearly one million inhabitants, substantial transit activity, and a mixture of formal and informal wastewater and decentralized sanitation systems (Figure 1C).^22^ The study sites were distributed across a branching canal network in which two upstream arms merged into a downstream channel (Figure 1A, B). The high background diversity in untreated wastewater from human, animal, plant, environmental and sewage-resident microbial input can mask low-abundance pathogens, especially viruses.^23–26^ To enhance sensitivity for viral detection, we performed hybrid-capture enrichment (Illumina VSP V2) on all samples, generating a median of 7.8×10⁶ reads per sample (Supplementary figure 1A). We used two parallel metagenomic classification approaches to ensure sensitive pathogen detection while minimizing false positives. We performed taxonomic classification with KrakenUniq (KU) using three unique k-mer thresholds: a relaxed discovery threshold, a sequencing-depth-normalised threshold and a stringent threshold to minimise false positives while preserving recall.^27,28^ We also used the read-mapping-based EsViritu (ESV) pipeline, which controls for false positives using a breadth-of-coverage threshold (See Methods).

**Figure 1.**
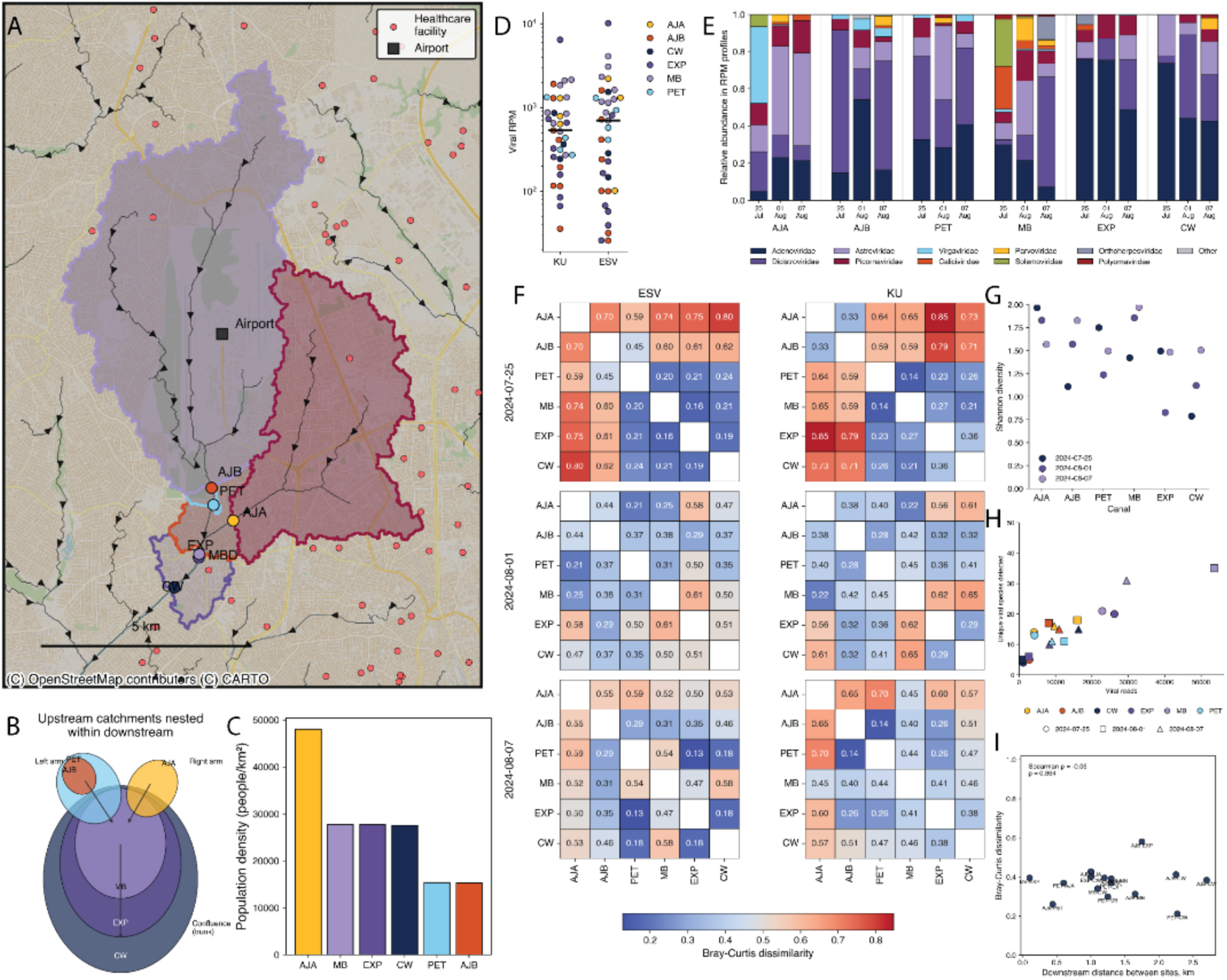
a) Sampling map of Oshodi/Isolo in Lagos, with sampling sites and catchment areas annotated. Arrows indicate drainage, and direction of flow. Canal sampling points: Ajibade Babatola (AJB), Petrocam Canoe (PET), Ajao Estate (AJA), Mass Burial (MB), an Extra Point (EXP) and Canal Water (CW). b) Nested catchment areas mapped by canal c) Population density for each catchment, calculated from population statistics obtained from es.world. d) Viral read per million reads assigned by KrakenUniq (KU) and Esviritu (ESV) respectively across samples, coloured by sampling site. e) Comparison of relative abundance of viral families across all three time points for each sampling site in the canal annotated at the bottom, as assigned by ESV. f) Bray-Curtis dissimilaries between viral family-level relative abundance profiles across the six sampling sites for each sampling date by Esviritu (ESV) and KrakenUniq (KU) taxonomic assignment. g) Comparison of Shannon diversity across the six sampling sites for each collection date. h) Comparison of unique viral species detected by ESV to the number of assigned viral reads. Datapoints are coloured by sampling site, and shaped by collection date. i) Distance decay of relative abundance profiles assigned by ESV between hydrologically connected canal pairs.

### Wastewater virome composition reflects sampling network relationships

Despite enrichment, the proportion of viral reads remained very low: ranging from 0.005% to 1% across samples and methods, with a median of 0.05% (Figure 1D, Supplementary figure 1B).^23,29–32^ Rarefaction analysis showed that unique viral species detections had not saturated (Supplementary figure 1C), indicating that additional sampling or deeper sequencing would likely recover further diversity as expected. Both approaches nonetheless detected a diverse virome spanning vertebrate, plant, microbial and environmental viral families (Figure 1E, Supplementary figure 1D). KU (relaxed threshold) identified 20 viral families, 84 genera and 88 species, while ESV identified 11 families, 18 genera and 38 species after breadth-of-coverage filtering. Bacteriophages comprised over 80% of viral species detected for both methods, comparable to estimates from recent wastewater virome studies.^9,21,29,33^ Overall, although KU detected more than twice as many viral species as ESV, both methods classified comparable per-sample viral read abundances with no significant difference across samples (Figure 1D). This indicates that KU’s additional taxa were each represented by relatively few reads. Together with the non-saturating rarefaction curves, this suggests that deeper sequencing would enable more comprehensive characterization of virome composition.

To inform the design of an optimal surveillance framework, we investigated whether a single downstream site could capture the virome composition of the whole canal network, or whether distributed sampling was necessary (Figure 1A). Towards this, we investigated the spatial structure of virome composition across the canal network using family-level Bray-Curtis dissimilarities across sites, dates and methods. We found that pairwise dissimilarity was highest on the first sampling date for both classifiers (Figure 1F). Compositional differences were driven by shifts in *Adenoviridae* and *Astroviridae* across methods, with *Dicistroviridae* (ESV) and *Picornaviridae* (KU) also contributing (Figure 1E, Supplementary figure 1D). Shannon diversity did not differ across sampling sites (Kruskal-Wallis=0.17) or dates (p=0.50, all BH-corrected p > 0.34). This indicates that compositional differences were not accompanied by detectable differences in overall alpha diversity (Figure 1G). Richness differed across canals (p=0.04) but not after correcting for read depth (p=0.13). Read depth did not differ significantly across canals (p=0.19). As expected, richness was strongly correlated with read depth at the sample level (Pearson r =0.84, p < 0.0001) (Figure 1H).

To further inform the potential sampling distribution along the canal network, we performed distance-decay analyses on all site pairs across methods to test whether taxonomic similarity decayed downstream along the direction of flow. A distance-decay relationship was detectable only in ESV profiles at the first date for hydrologically connected sites (Mantel p=0.07, Spearman p=0.033) (Figure 1A-B, F, I, Supplementary figure 1E). On that date, AJA was the primary compositional outlier while PET, MB, EXP and CW formed a cohesive cluster, indicating confluence composition reflected predominantly left-arm input (Figure 1A-B). This structure was absent on other dates and for the KU analyses. Similarity was independent of hydrological connectivity, suggesting connectivity’s influence was transient or method-dependent rather than a stable network feature (Figure 1). As the terminal site CW had <1000 viral reads on two of the three dates, we repeated these analyses excluding those samples. This strengthened the relationship on the first date alone (ρ =-0.73, p=0.040).

To further test whether downstream sites can serve as proxies for upstream sites, we assessed whether upstream viral communities form nested subsets of downstream communities. Towards this, we partitioned taxonomic dissimilarity into turnover and nestedness components with the Baselga-Sørensen framework. At family level, dissimilarity was nestedness-dominated on all three dates for both methods (nestedness 56-89%), but this did not persist at species level ( Supplementary Figure 1F). As most viral families were present at most sites, family-level nestedness largely reflected shared family membership rather than true subset structure. At species level, where the sparser matrix improved the power of the test, dissimilarity was turnover-dominated on the date with most even sequencing depth (2024-08-07, 79%) and more evenly partitioned on the others (nestedness 43-54%). Canals thus shared viral families but often differed in the underlying classified species, consistent with taxonomic replacement rather than nested loss. Accordingly, we found no significant nestedness at species level for either classifier: network-level NODF did not exceed a prevalence-preserving null on any date (ESV p =0.57-0.93; KU p=0.09-0.98). Directional containment of upstream species downstream was low (mean 0.41-0.47), and species richness was not positively related to flow position on any date (ρ=−0.66 to −0.09, all p>0.15) (Supplementary figure 1G). Species richness is strongly depth-dependent, however, and our rarefaction curves had not saturated (Supplementary Figure 1C), so these species-level estimates represent depth-limited lower bounds. On 2024-08-01, two canals fell below a viral read threshold of 1000, limiting inference for that date. The one directional signal consistent across both resolutions and classifiers was the relative isolation of AJA: its containment in the downstream confluence was lowest on 2024-07-25 (0.36 ESV, 0.09 KU) and rose thereafter, consistent with AJA’s isolation in the distance-decay analysis. Overall, the wastewater virome showed no statistically supported nestedness along the canal network and was better described by taxonomic turnover among sites.

Taken together, these analyses suggest that virome composition across this canal network was shaped more by transient, local inputs than by stable downstream flow. For surveillance, this suggests that we cannot rely on a single terminal site to integrate upstream viral signals. Distributed sampling across the network would more reliably capture pathogens of public-health concern.

### Diverse pathogenic viruses are present in Lagos wastewater

To characterize the microbial populations relevant to public health, we filtered our results to viruses known to infect humans. KU and ESV identified 15 and 16 human-associated viral species from eight and seven families respectively. As read abundance should not be interpreted as a direct proxy for pathogen abundance, particularly after target enrichment, we focused on presence-absence patterns and interpreted relative abundance only within the compositional constraints of metagenomic data.^34^ Across samples and methods, the dominant human-associated viral families were the common enteric *Adenoviridae* (median 30% of viral reads across samples), *Astroviridae* (12%) and *Picornaviridae* (11%), with *Caliciviridae* (4.3%), *Orthoherpesviridae* (3%), *Parvoviridae* (2%) and *Polyomaviridae* (1%) detected at lower abundance (Figure 2A), consistent with previous studies of Nigeria’s sewage virome.^9,29^ Family-level composition was consistent between methods, although KU failed to detect any viruses in several low viral read samples after k-mer thresholding, which ESV retained after breadth-of-coverage filtering (Figure 2A-B). In particular, ESV estimated higher relative abundances of *Parvoviridae* and *Caliciviridae,* while KU results indicated higher abundance of *Picornaviridae* reads. At genus level, ESV identified diverse viruses that were not detected by KU, such as sapovirus, adeno-associated virus and enterovirus (Figure 2B). No viral species or family showed a significant change in abundance or detection frequency across the three sampling weeks after correction for multiple testing (all q > 0.15, Figure 2C). Additionally, per-sample species richness showed no significant temporal trend (Page trend test p=0.10, Friedman p= 0.18). *Parvoviridae*, driven by adeno-associated virus 2, showed a suggestive increase in detection over time, but this did not survive correction.

**Figure 2.**
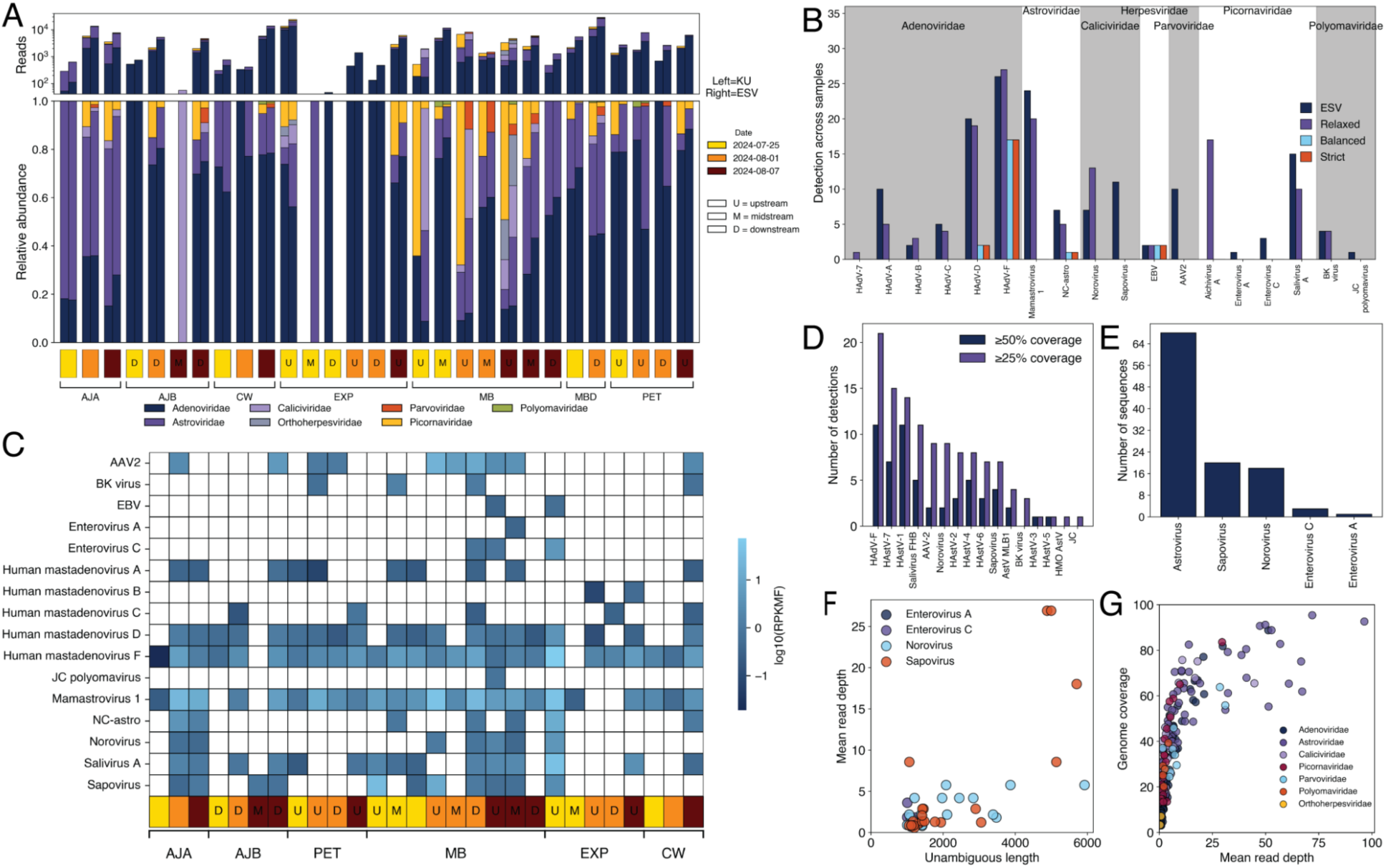
a) Relative abundance and absolute reads counts assigned to viral families, annotated in colour as per legend, by KU (left grouped bar) and ESV (right grouped bar) across all samples. Samples are grouped by canal as per text annotation, and annotated by date and sampling position if available in colour and text as per legend. Ajibade Babatola (AJB), Petrocam Canoe (PET), Ajao Estate (AJA), Mass Burial (MB), an Extra Point (EXP) and Canal Water (CW). b) Number of detections of viruses across all samples by different approaches in this study. BC represents the breadth-of-coverage filtered results from ESV. Relaxed, balanced and strict represent the results filtered by the respective unique k-mer thresholds of KU stipulated in Methods. c) Reads per kilobase of transcript per million filtered reads (RPKMF) for viruses of clinical concern across all samples, assigned by ESV. Samples are grouped by canal as per text annotation, and annotated by date and sampling position if available in colour and text as per legend in A. d) Number of detection for viruses of clinical concern across all samples, with detection threshold defined at 25% and 50% genome coverage as per legend. e) The number of contigs recovered by ESV for viruses of clinical concern across samples. f) The relationship between unambiguous genome length (nt) and mean read depth for contigs recovered by ESV for viruses indicated by colour as per legend. g) The relationship between genome coverage and mean read depth for all sequences recovered by ESV, annotated for viral families as per legend.

Several pathogens, including noroviruses and adenoviruses, were consistently detected across methods and thresholds. Others, such as enteroviruses, were less concordant (Figure 2B, C). As expected from untreated wastewater, a small number of enteric species dominated (Figure 2A, C). *Mastadenovirus* was the most abundant genus in both methods, and was detected across all sites, time points, methods and thresholds. This was driven by Human mastadenovirus F (HAdV-F), including the common HAdV41, a leading cause of viral gastroenteritis and diarrhoea in children globally.^35–38^ Human mastadenovirus A-D were detected across multiple samples under the KU relaxed and ESV breadth-of-coverage thresholds, but only HAdV-D passed the depth-normalised k-mer thresholds (Figure 2B). *Mamastrovirus* was the second most frequently detected genus, largely attributable to Mamastrovirus 1 (HAstV1), the dominant human astrovirus serotype worldwide.^6,9,29^ HAstV1 was detected in over 20 samples by both methods across all sites and time points. Although none of the HAstV1 detections passed the depth-normalised KU thresholds, ESV recovered several HAstV1 contigs above 50% genome coverage.

Noroviruses (*Caliciviridae*) were detected across multiple samples from all sites and time points under both the relaxed KU and ESV breadth-of-coverage thresholds (Figure 2B, C). KU read support across samples was moderate, with moderate unique k-mer support and database k-mer space coverage, consistent with low-abundance detection in complex samples (Supplementary figure 2A-B). Although no detections passed the depth-normalised KU thresholds, seven samples passed the ESV breadth-of-coverage threshold and generated 18 contigs with unambiguous lengths above 1000 nt (Figure 2B, D, H, Supplementary figure 2C). ESV recovered 11 norovirus contigs, including seven Genogroup II (GII) detections and three GI detections, as well as one GIV detection.^39^ Genotype detections were primarily associated with GII.17 (n=4), GII.2 (n=3), and GII.3 (n=2), among the most globally prevalent GII genotypes, and GI.3 (n=2), the most globally prevalent GI genotype.^39^ Mean read depth was often low despite moderate contig lengths, limiting suitability for detailed genomic epidemiological analyses and confidence in genotype assignments (Figure 2D, F-G).

This limitation applied across most detected pathogens (Figure 2F-G). After filtering for human viruses, a median of only three viral species per sample reached >50% genome coverage, with read depth generally insufficient for robust downstream analyses including phylogenetic reconstruction. Nevertheless, we recovered 57 sequences at >50% coverage across HAdV-F, noroviruses, sapoviruses, several human astroviruses and saliviruses (Figure 2D, F-G). Lowering the threshold to >25% coverage expanded both sequence recovery and species range (Figure 2D). Applying a joint threshold of 50% coverage and a mean read depth of 10 yielded 42 sequences - mostly human astroviruses and HAdV-F - which may be suitable for genomic epidemiology (Supplementary Figure 2D).

*Enterovirus* A, *Enterovirus* C and *Sapovirus* failed to meet all KU k-mer thresholds (Figure 2B). ESV detected *Enterovirus* A and *Enterovirus* C above the breadth-of-coverage threshold in one and three samples respectively. Enterovirus C read counts and coverage were lower than noroviruses, reaching a maximum of 17% genome coverage (Figure 2F, Supplementary figure 2D). ESV recovered contigs exceeding 1000 unambiguous nucleotides from three samples, all with low mean read depths ranging from one to four (Figure 2F-G). Enterovirus A was detected in one sample with limited read support and 15% genome coverage, yielding a consensus sequence at a mean read depth of one. KU assigned reads to Enterovirus A or C in 15 samples, but all detections showed very limited unique k-mer support, low database k-mer space coverage and elevated k-mer duplication, consistent with repeated sampling of a small genomic region rather than broader genome representation (Supplementary figure 2E). Sapoviruses were detected in 11 samples after ESV breadth-of-coverage filtering (Figure 2B), with genomic coverage up to 75% (Figure 2E-G). ESV generated 20 contigs exceeding 1000 unambiguous nucleotides at relatively high mean read depth (Figure 2F-G, Supplementary figure 2F). Genotyping suggested an approximately even GI/GII distribution, but given the uncertainty of genogroup assignment without phylogenetic confirmation we report these conservatively at species level. KU also assigned reads to Sapovirus, but unique k-mer support was consistently low, with a maximum of approximately 350 unique k-mers (Supplementary figure 2F).

Together, these findings indicate that pathogenic enteric viruses are widespread in Lagos wastewater. However, many detections were low abundance and could not be confidently resolved across methods at the available sequencing depth. This underscores a key limitation of metagenomic wastewater surveillance: true low level detections must be distinguished from false positives using conservative thresholds. However, the recovery of higher-coverage contigs for some pathogens failing depth-normalized unique k-mer thresholds suggests that these thresholds may be overly stringent in some contexts.

### Diverse bacterial pathogens are detectable in Lagos wastewater

Although our approach aimed to specifically enrich for viruses, the majority of reads were assigned to bacterial species, including known human pathogens (Supplementary Figure 1B). Our sampling coincided with a major cholera outbreak in Nigeria, with more than 4200 suspected cases in June to July.^40^ This provided an opportunity to assess whether *Vibrio cholerae* and other bacterial pathogens could be detected by our wastewater metagenomic protocol, identifying candidates for future targeted surveillance panels. For our bacterial analyses, we used KU with the same thresholding approach as for the virome. For cross-validation with breadth-of-coverage thresholding, we applied the approach developed by Crits-Christoph *et al* as ESV was developed for viruses.^41^ KU identified a highly diverse bacterial community of 291 families, 762 genera and 1888 species, spanning human microbiome, fecal, environmental and aquatic taxa.^25^ After filtering to genera containing potentially human pathogenic species, we identified 42 families, 68 genera and 139 species across samples (ranging 13-65 per sample) (Figure 3A). Despite this diversity, a small number of genera dominated, including ubiquitous environmental, human microbiome and fecal-associated genera such as *Pseudomonas* (mean 2.5% of all bacterial reads across samples), *Aliarcobacter* (2.3%), *Aeromonas* (0.67%), *Klebsiella* (0.2%), *Escherichia*, and *Streptococcus* (0.5%) (Figure 3A). At species-level, we detected emerging zoonotic (e.g. *Streptococcus suis*) and enteric pathogens (*Salmonella and Shigella spp.*), opportunistic pathogens (*e.g. Klebsiella pneumoniae*), and pathogens of public health concern (e.g. *Vibrio cholerae, Bordetella parapertussis*) (Figure 3A, B). However, detection consistency varied across approaches, with many species-level detections failing to replicate across increasingly stringent unique k-mer thresholds, breadth-of-coverage analyses and metagenome assembly (Figure 3B).

**Figure 3.**
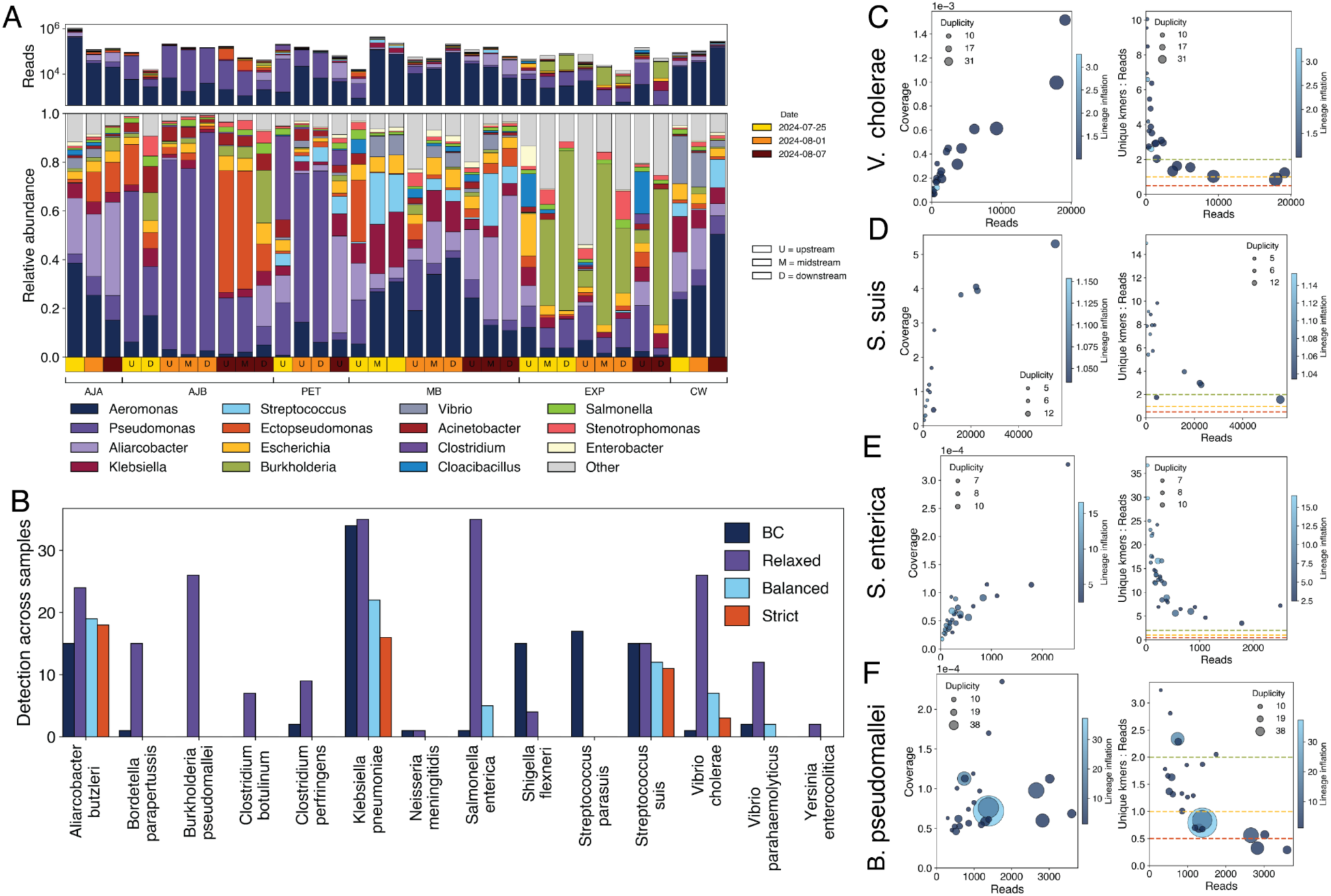
a) Relative abundance and absolute reads counts assigned to bacterial genera, annotated in colour as per legend, by KU across all samples. Samples are grouped by canal as per text annotation, and annotated by date and sampling position if available in colour and text as per legend. Ajibade Babatola (AJB), Petrocam Canoe (PET), Ajao Estate (AJA), Mass Burial (MB), an Extra Point (EXP) and Canal Water (CW) b) Number of detections of bacterial pathogens across all samples by different approaches in this study. BC represents the breadth-of-coverage filtered results. Relaxed, balanced and strict represent the results filtered by the respective unique k-mer thresholds of KU stipulated in Methods. c1) Relationship between k-mer coverage (coverage of the database k-mer space) and assigned read counts for *V. Cholerae*, as per KU. Points are coloured by lineage inflation, defined as the ratio of total clade-level reads to reads assigned directly to the species node, and sized by k-mer duplicity, the multiplicity with which the same k-mers are sampled (or duplicated). c2) The relationship between the ratio of unique k-mers to reads counts and the number of reads for *V. Cholerae*, as per KU. Points are coloured by lineage inflation, and sized by k-mer duplicity. d1) Relationship between k-mer coverage and assigned read counts for *S.suis*. d2) The relationship between the ratio of unique k-mers to reads counts and the number of reads for *S.suis*. e1) Relationship between k-mer coverage and assigned read counts for *S. enterica*. e2) The relationship between the ratio of unique k-mers to reads counts and the number of reads for *S. enterica*. f1) Relationship between k-mer coverage and assigned read counts for *B. pseudomallei*. f2) The relationship between the ratio of unique k-mers to reads counts and the number of reads for *B. pseudomallei*.

*V. cholerae* was detected in 26 samples across all canals and time points under the relaxed KU k-mer threshold (Figure 3B, C). As reference-based metagenomic approaches can be biased by sequence similarity among closely related taxa and many *Vibrio* species are naturally abundant in aquatic systems (Figure 3B), we validated detections using more stringent thresholding approaches, breadth-of-coverage analyses, and metagenome assembly. Seven and three samples passed the balanced and strict thresholds respectively, and only one sample was confirmed by breadth-of-coverage analyses (Figure 3B). In the KU analyses, lineage inflation, a measure of species assignment overrepresentation, was generally low, indicating reads were assigned directly to the *V. cholerae* node rather than accumulating across the genus, and several samples showed strong unique k-mer support (Figure 3C). Although database k-mer space coverage was low, consistent with sparse genome representation, the combination of repeat threshold-passing detections, low lineage inflation, breadth-of-coverage confirmation and assembly of ten contigs (533-2352 nt) supports true and recurrent detection of *V. cholerae* during the 2024 outbreak.

We detected other bacterial pathogens of concern with varying levels of support (Figure 3B). The most consistent was for *S. suis* (Figure 3B, D). Most *S. suis* detections passed multiple thresholds, were confirmed by breadth-of-coverage, and showed low lineage inflation and strong unique k-mer support across samples. Other pathogens showed consistent signals but warrant cautious species-level interpretation. For example, *Salmonella enterica* was detected in multiple samples, with five passing the KU balanced threshold and one confirmed by breadth-of-coverage, though moderate lineage inflation and low k-mer coverage warrant cautious species-level interpretation (Figure 3E). Similarly, *Shigella flexneri* was detected in four samples and, despite failing stringent thresholds, showed low lineage inflation, strong unique k-mer support and comparatively high database k-mer coverage. Unlike the relatively consistent *V. cholerae* and *S. suis* detections, several other bacterial pathogens showed weaker agreement across thresholds and analyses. For example, we detected *Burkholderia pseudomallei*, the environmental agent of melioidosis, under relaxed thresholds in multiple samples, several with high read counts (Figure 3B, F). Melioidosis is thought to be endemic in Nigeria, with an estimated 13 000 cases annually and seropositivity of up to 30% reported in an Ebonyi State cohort.^42–45^ However, no detections passed the balanced or strict k-mer thresholds and database k-mer coverage was extremely low, consistent with low abundance, reads concentrated in limited genomic regions and/or conserved sequences shared with related environmental taxa (Figure 3F). Assembly yielded four contigs (503-1585 bp) assigned to *B. pseudomallei* by BLAST. Detection of *Burkholderia pseudomallei* is therefore plausible given its environmental reservoir and suggested endemicity in Nigeria, but species-level resolution will require bacterial-optimised workflows, cross-sample co-assembly, confirmation by PCR and long-read approaches.^46,47^

Collectively, these findings support the presence of genuine bacterial pathogen signals in Lagos wastewater, including *V. cholerae.* It also highlights the importance of complementary confirmation approaches to resolve the taxonomic ambiguity inherent to reference-based metagenomics, particularly for bacterial pathogens with shared, conserved or low-complexity genomic regions.

### Antimicrobial resistance genes are abundant in Lagos wastewater

Global sewage-based resistome surveys have reported a high total AMR load for Nigeria.^6,7,48^ To assess whether our sequencing approach could support AMR monitoring, we investigated the diversity and abundance of antimicrobial resistance genes (ARGs) within our dataset using ARGprofiler against the PanRes database, which includes six major ARG databases. Our catchment areas include three major hospitals ( in AJA, AJB and CW) and several primary and secondary healthcare facilities that may contribute to wastewater AMR burden alongside environmental and agricultural sources (Figure 1A). We found that 0.02%-1.8% of reads mapped to ARGs across samples (Figure 4A). We identified 1110 ARGs in total, corresponding to 281 unique ARGs and 1.2 million fragments (Figure 4 A-C). The most abundant ARGs conferred resistance to drug class aminoglycoside (277 ARGs), followed by beta-lactam (264) and fluoroquinolone (134) (Figure 4C, D), consistent with previous studies.^6,7,48^ These classes include higher priority agents listed among the critically important antimicrobials for human use by the WHO.^49^ ARG richness and abundance patterns differed across classes: sulfonamide ARGs had higher RPKM than aminoglycoside and beta-lactam ARGs (figure 4D, E). This suggests that aminoglycoside and beta-lactam resistance were represented by a broader diversity of ARGs, whereas sulfonamide resistance was driven by fewer but more abundant genes.

**Figure 4.**
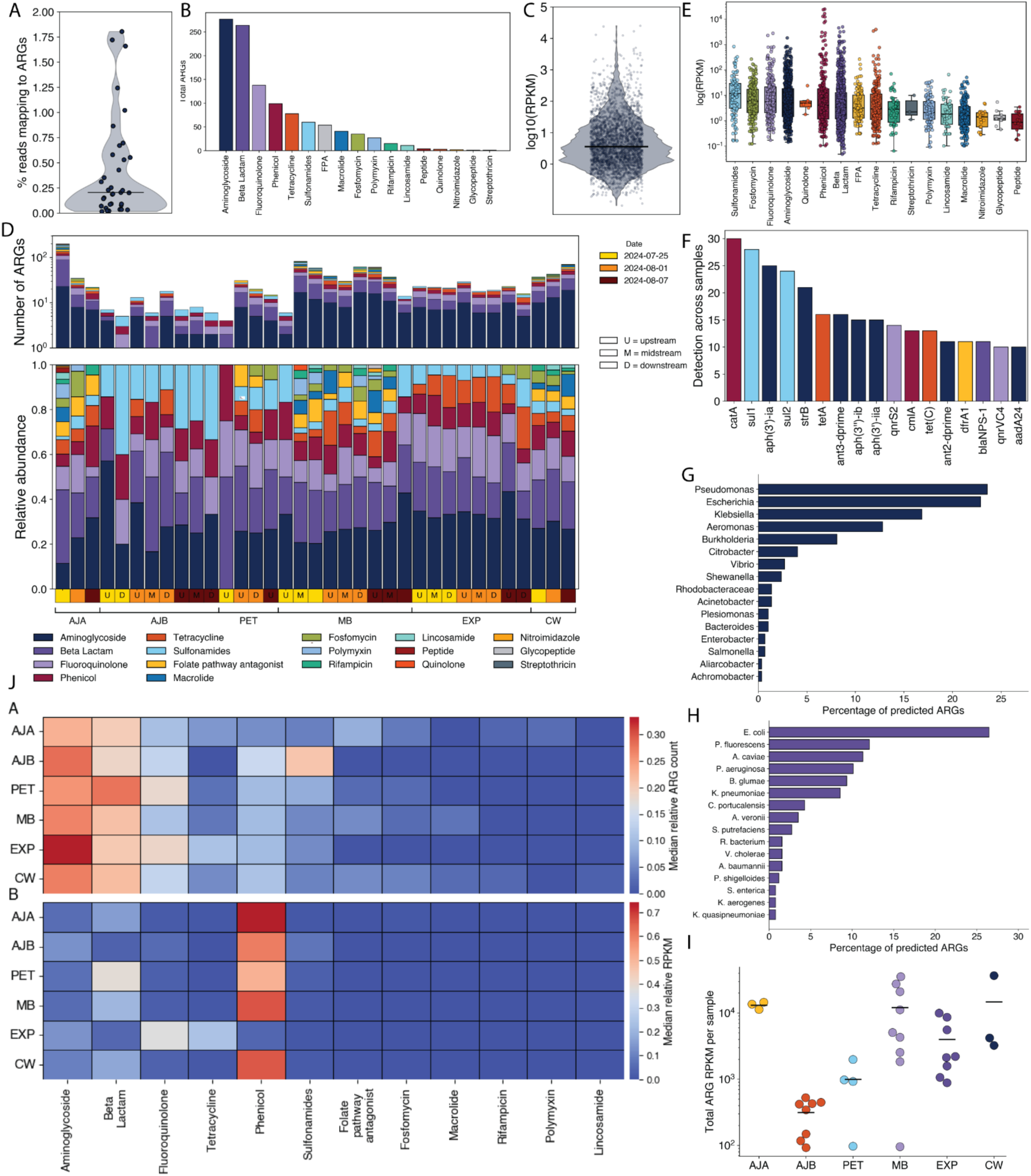
a) Percentage of reads mapping to ARGs in the PanRes database across samples. b) Total number of ARGs by resistance drug classes. c) Overall reads per kilo base per million mapped reads (RPKM), used to normalise ARG read counts by gene length and sequencing depth. d) Relative abundances of ARGs aggregated on their resistance drug classes as per legend below. Samples are grouped by canal as per text annotation, and annotated by date and sampling position if available in colour and text as per legend. Ajibade Babatola (AJB), Petrocam Canoe (PET), Ajao Estate (AJA), Mass Burial (MB), an Extra Point (EXP) and Canal Water (CW) e) Distribution of RPKM mapping to ARGs across different resistance drug classes. f) Detection of canonical ARGs across samples, coloured by resistance drug classes as per legend in D. g) Percentage of ARGs with predicted taxonomic origin that belong to bacterial genera h) Percentage of ARGs with predicted taxonomic origin that belong to bacterial species. Only the top 15 predicted species are shown, representing 96% of assigned ARGs. i) Total ARG RPKM per sample across all canals.

Fifteen ARGs were ubiquitous, detected in more than 50% of samples (Figure 4F). The most abundant ARGs were non-canonical genes associated with fluoroquinolone, tetracycline and beta-lactam resistance. The most abundant canonical ARGs were *catA*, a chloramphenicol acetyltransferase associated with phenicol resistance, and *sul1*, a sulfonamide resistance gene reported on small plasmids across diverse bacterial species in Africa (Figure 3B).^7^ This is followed by *APH(3’)-Ia* (aminoglycoside phosphotransferas), *sul2, strB* (aminoglycoside phosphotransferase) and *tetA* and C *(tetracycline efflux-associated*). The high prevalence of *strB*, *tetA* and *tetC* is consistent with previous studies of the Nigerian resistome.^6^ We detected two mobile colistin resistance gene (*mcr)* across four samples.^50^

Our short-read data have limited power to resolve the bacterial host, genomic context and mobility of these ARGs. To address these questions within the constraints of short-read data, we used the CARD reference-based k-mer classifier implemented in RGI, which does not depend on assembly or flanking information.^51^ The taxonomic origin of 55% of sequences could not be predicted. Among classified ARGs, most were assigned to *Pseudomonas* (24%), *Escherichia* (23%), *Klebsiella* (17%), *Aeromonas* (13%) and *Burkholderia* (8%) (Figure 4G). At species level, the majority of classified ARGs were predicted to originate from *Escherichia coli* (26%, chromosomal or plasmid context), followed by *Pseudomonas fluorescens* (12%), *Aeromonas caviae* (11%), *P. aeruginosa* (10%) and *Burkholderia glumae* (9%, chromosomal) (Figure 4H). Of the ARGs with predicted taxonomic associations, 7% mapped specifically to a plasmid context and 26% to a chromosomal context. Only one ARG was assigned to a non-*Klebsiella* plasmid, predicted to originate from *Aeromonas cavia*e. Among the chromosomally associated ARGs, 32% were predicted to originate from *Burkholderia glumae*, 32% from *Pseudomonas* spp. and 16% from *Aeromonas* spp. Four ARGs were assigned to *Vibrio cholerae* with unknown genomic context.

We compared ARG abundance and diversity across catchment areas in an exploratory analysis. Per-sample ARG abundance differed significantly across canals (Kruskal-Wallis p =0.0008). CW, AJA and MB had the highest abundance, but were statistically indistinguishable from one another (Figure 4I). AJB had significantly lower ARG abundance than all canals excluding PET (Dunn’s test, BH-corrected q < 0.05). Overall, the resistome composition was broadly consistent across canals by ARG count. Aminoglycoside and beta-lactam ARGs accounted for the largest share of distinct ARGs in every canal (23-33% and 19-28%), followed by fluoroquinolone (Figure 4JA). No canal was a clear compositional outlier by ARG count, excepting a significantly higher relative sulfonamides ARG count for AJB. However, abundance-weighted composition diverged sharply from ARG count. In most canals, phenicol ARGs dominated by RPKM (51-74%) despite representing only 6-14% of distinct ARGs, reflecting a small number of highly abundant ARGs (Figure 4Jb). EXP was the exception: phenicol abundance was low, and fluoroquinolone and tetracycline dominated instead, again exceeding their proportion by ARG count. In both cases, abundance was concentrated in a few genes rather than a broader set of ARGs.

Overall, we observed a diverse and abundant ARG repertoire across all sampling locations, dominated by aminoglycoside, beta-lactam and fluoroquinolone resistance including clinically important genes such as *mcr* and *sul1*. This supports the approach for AMR surveillance in Nigeria, although short-read data limit resolution of the bacterial hosts, genomic context and mobility of these ARGs - which future long-read sequencing should address.^46,52^

## Discussion

We demonstrate that sequencing of untreated wastewater from open drainage enables community-level monitoring of diverse circulating pathogens and AMR markers. Accurate community-level profiling of pathogen diversity is necessary to guide surveillance efforts, including prioritising of pathogens for targeted assays and amplicon sequencing for higher genome coverage, genotyping and variant tracking.^30^ Using virus-enriched metagenomic sequencing, we identified diverse human viral and bacterial pathogens as well as ARG profiles from the same samples, providing proof-of-concept for integrated virome, bacteriome and resistome surveillance for setting with limited sewage infrastructure.^6,7,9^ We showed consistent detection of the enteric pathogens that are major causes of the high diarrheal disease burden in Nigeria, where clinical surveillance is limited by healthcare-access disparities and diagnostic constraints.^53–55^ *V. cholerae* detection is particularly important for Lagos and Nigeria overall, as cholera risk has been closely tied to inadequate sanitation and climate-related disruption of water systems.^16–18,56^ The detection of animal-associated viruses, including diverse kobuviruses and rat hepatitis E virus alongside *Streptococcus suis*, supports the additional One Health value of our metagenomic surveillance approach. Wastewater metagenomics also enabled us to capture the resistome circulating across the wider population, including from animal and environmental sources deposited in open drainage.^6,57,58^ Wastewater AMR surveillance is especially valuable for African settings, where the AMR burden is high and regional ARG diversity is poorly characterised by sparse clinical surveillance.^7,59^

Our findings should be interpreted in the context of limitations in our sampling and sequencing strategies. Using our grab sampling approach, each sample provides a representation of shedding at a single timepoint, rather than a composite representation of all shedding over the course of the day, introducing variability in pathogen abundance per sample and limiting detection sensitivity.^60^ Despite target enrichment, our viral read proportions remained low. This likely reflects a combination of the dominant bacterial background and low viral input, degraded nucleic acids, PCR inhibitors and variable efficiency in viral concentration, purification and extraction protocols.^37,61,62^ Accordingly, viral detections had not saturated, and richness, diversity, and containment estimates therefore represent lower bounds.^30^ Most notably, respiratory viruses were not detected above discovery thresholds, even where probes were available.^23^ SARS-CoV-2 reads were detected in 7 and 11 samples by ESV and KU respectively, but none passed the lowest coverage and unique k-mer thresholds. RSV detection was expected, as RSV peak prevalence in Nigeria occurs during the rainy season (May to October).^63^ Influenza in Lagos shows irregular bi-annual peaks (in January-March and September-November), though cases occur year-round.^64^ Sampling open canals in July and August, during this lower-transmission period, rather than sewered wastewater during peak respiratory virus circulation, likely further contributed to the absence of detection.^64^ Overall, our approach is intended to maximize detection of priority viruses, rather than providing a proportional representation of the microbial community. As such, results should be interpreted as measures of presence and relative diversity rather than unbiased abundance. Notably, reference-based pathogen detection in untreated wastewater is challenging owing to high microbial background, short-read limitations, closely related environmental species and unevenly curated databases.^25,26,65^ Low-abundance detections may therefore reflect true positives, but could also represent database artefacts or method-specific misclassification.^27,28,30,66,67^ Robust validation of such detections requires genome-wide evidence and/or cross-validation. Consequently, we used genome-wide coverage thresholds, multiple taxonomic classification methods and depth-normalised thresholds. However, residual false-positives cannot be excluded. Discordance in our virome analyses between EsViritu and KrakenUniq may reflect differences between mapping-based versus k-mer-based detection, as well as database structure or threshold choice rather than true disagreement.^34,68,69^ The recovery of contigs with >25% genome coverage for some detections failing depth-normalised k-mer thresholds with KU suggests these thresholds may have been overly stringent. Further, this study provides a localized, time-restricted baseline rather than longitudinal surveillance, and does not fully capture temporal, seasonal or spatial variation in wastewater microbial communities across Lagos.^7,23,70^ Available sampling has limited power to recover of spatial structure from sequencing data, or characterize of site specific variability in microbial diversity.

Despite these limitations, wastewater metagenomic surveillance complements clinical systems by capturing population-level pathogen and AMR dynamics from human, animal and environmental sources in resource-limited settings. Lagos is a particularly strategic setting for developing regionally-tailored sentinel surveillance systems owing to its high population density, socioeconomic diversity, regional connectivity, international travel links and complex human-animal-environment interfaces.^16–18^ Moving forward, we propose a tiered wastewater surveillance model that combines periodic baseline metagenomic surveys with routine targeted qPCR and amplicon sequencing, supporting early warning of outbreaks, identification of transmission and AMR “hotspots”, and enabling prioritization of public health interventions.

## Methods

### Ethics

This study did not involve any human subjects. Approval to conduct the study was obtained from Lagos State Waste Management Office (LSWMO/OS&M/3274), Ikeja, Lagos.

### Sample collection

We collected untreated wastewater samples from a network of open drainage canals situated in Isolo Local Council Development Area (LCDA), in Oshodi/Isolo LGA. Oshodi/Isolo LGA is a major local government area in Lagos (estimated population of 931 000) which serves both as a transit hub and a residential area. We selected Oshodi/Isolo LGA for this study owing to its high population density, socioeconomic diversity that broadly reflects Lagos, potential value as a sentinel surveillance site due to its role as a major transport hub and the coexistence of formal and informal sewage networks. The area has limited formal sewage infrastructure, but some households use systems such as soakaways or septic tanks connected to individual buildings. Wastewater in this setting receives direct inputs from household latrines, vendors in markets and open drainage systems among other sources. Using the WHO GIS web tool ES.World 4.0, we characterised wastewater catchments for each potential sampling site, including estimated surrounding population size, building footprints, natural and man-made drainage networks and roads. Sampling sites were selected based on accessibility, wastewater flow and proximity to large catchment populations or sewersheds exceeding 50 000 people. The sites include: Ajibade Babatola (AJB), Petrocam Canoe (PET), Ajao Estate (AJA), Mass Burial (MB), an Extra Point (EXP) and Canal Water (CW). The sites spanned a branching canal network: the left arm included AJB and PET, spanning 2.7 km from upstream to terminal downstream sites, while the right arm included AJA, spanning 2.2 km to CW (See Figure 1). After confluence, sites were arranged sequentially as MB, EXP and CW, with CW representing the terminal site integrating upstream inputs (Figure 1A, B). We collected a total of 38 untreated wastewater samples in the early morning using the grab sampling method over a period of three weeks, from 25 July 2024 to 7 August 2024. Collection was done from multiple points at each location (upstream (UPS), midstream (MDS) and downstream (DWS)), with the coordinates of the locations recorded using GPS. We collected 500 mL of untreated wastewater in prelabelled sample bottles, transported samples to the laboratory in cooler boxes within 2 hours of collection and stored them at 4°C until processing.

### Sample processing and nucleotide extraction

We performed particle concentration for the samples using 7.5% w/v of polyethylene glycol (PEG) and 1.7% w/v of Sodium Chloride (NaCl) in 40ml of the sample. The mixture was subsequently centrifuged at 1200g and 4oC for two hours, with the pellet resuspended in 500 µl of phosphate buffer saline (PBS). Resuspensions were transferred to cryotubes and either processed immediately or stored at −80 °C until nucleic acid extraction. Nucleic acids were extracted from 200 µL of the sample using the QIAamp Viral RNA Mini Kit (Qiagen) according to the manufacturer’s protocol, with elution in 50/60 µL.

### Target enrichment, library preparation and sequencing

Untreated wastewater contains complex inputs from human, animal, plant, environmental and sewage-resident microbial communities.^25,26^ This high background diversity can mask low-abundance pathogens, especially viral targets. Additionally, the sewage virome is dominated by bacteriophages and plant viruses.^23,24^ Wastewater-based metagenomic viral surveillance therefore requires target enrichment to substantially improve sensitivity for detecting low-abundance viral pathogens of clinical concern within highly complex sample backgrounds. To enhance detection sensitivity, we performed hybrid-capture target enrichment on all samples using the Viral Surveillance Panel V2, following the Illumina RNA Prep with Enrichment (tagmentation) protocol. The libraries were pooled and normalised to a starting concentration of 2nM and loaded in 20µl on an Illumina NextSeq 2000 system using a P3 300 cycle cartridge for 2x150bp reads, following the NextSeq workflow according to the manufacturer’s guidelines.

### Sequence data and taxonomic classification analyses

Raw FASTQ reads were trimmed and filtered with BBDuk to remove adapters, low-quality bases, short reads, low-complexity reads and reads with low average quality before downstream analysis.^71^ Negative extraction and no-template controls were free of target taxa, with background limited to known reagent-associated contaminants.^72^

Inconsistent or low-abundance detections in metagenomic sequencing of complex samples require conservative interpretation and targeted confirmation, underscoring the need for stringent false-positive control and multiple lines of evidence when applying reference-based metagenomics to untreated wastewater. Accordingly, we performed taxonomic classification using two approaches to account for possible false-positives. In the first approach (for both viruses and bacteria), we performed taxonomic classification with KrakenUniq v1.0.4 against the standard database containing the archaea, bacteria, viral, human and UniVec_Core reference sets. KrakenUniq (KU) combines Kraken’s rapid k-mer-based classification with efficient assessment of unique k-mer coverage for each species detected in a dataset. This approach improves recall and precision relative to other methods and can help distinguish low-abundance pathogens from false-positive assignments in infectious disease samples.^28^ For these analyses, we used three thresholds based on unique k-mer counts to control the false-positive rate while optimising recall: 1) a relaxed threshold for discovery, designed to maximise sensitivity while reducing background. A threshold of 1000 unique k-mers is recommended for KU^28^. However, this guidance was developed for reads of length up to 250 bp. As each assigned read is expected to contribute approximately L-k unique k-mers, where L is read length and k is k-mer length, shorter or more fragmented wastewater reads are expected to yield fewer unique k-mers per read. We therefore scaled the unique k-mer threshold to the average read length of the wastewater data, while ensuring that the k-mer threshold remained several-fold higher than the read-count threshold. Our relaxed threshold there was 700 unique k-mers. 2) a sequencing-depth-normalised balanced threshold optimised for recall (Balanced). As there is a positive relationship between sequencing depth and misclassified reads for k-mer-based methods, higher thresholds are needed at greater sequencing depths to control false-positive rates. Our balanced threshold reflects that by scaling the threshold to 700 unique k-mers per million sequencing reads ^69^ 3) a stringent sequencing-depth-normalised threshold designed to minimise false positives (Stringent) KU recommends approximately 2000 unique k-mers per million reads as the optimal unique k-mer threshold. Our stringent threshold scales this to fit wastewater data, with a threshold of 1300 unique kmers per per million reads.^27,28^ For KU, we defined lineage inflation as the ratio of total clade-level reads to reads assigned directly to the species node to the parent node. We note that k-mer coverage represents coverage of the database k-mer space, and high k-mer duplication values suggest repeated sampling of the same subset of k-mers rather than progressive expansion of genome-wide representation. Across methods, reads were normalized to read counts per million (RPM) for comparison.

To cross-validate our viral analyses, we also applied the read-mapping based EsViritu (ESV) pipeline (v1.2.1, with default settings, database v3.2.4) to validate detections and compare relative abundance across methods.23 In this approach, we used breadth-of-coverage (i.e. genome coverage) as the primary criterion for controlling false positives, as the even distribution of reads across a target reference genome is robust evidence of a true positive.41 Briefly, reads were mapped against the full database v2.0.2 using minimap273, after which CoverM (https://github.com/wwood/CoverM) was used to retain alignments meeting ≥90% average identity over ≥90% of the read length. For detection, we used a breadth of genome coverage threshold of 1000 nt as per the Esviritu recommendation.23 The Esviritu pipeline calculates metric RPKMF calculated as (Reads Per Kilobase of reference genome)/(Million reads passing Filtering).23 ESV additionally generated contigs, for which we used a detection threshold of an unambiguous sequence length of 1000 nt. All sequences were screened for human pathogens of interest by VirBot, and confirmed by BLAST.74

For bacteria, we used KU with the same thresholding approach as for the virome. For cross-validation with breadth-of-coverage thresholding for bacteria, to approximate the breadth-coverage approach of ESV, we applied the breadth-coverage approach developed by Crits-Christoph et al.^41^ Briefly, following quality trimming with BBDuk, reads were mapped to a custom database build from all genera with known human pathogens. Each mapped read pair was counted as a single observation. Only detections with a minimum coverage breadth of 500 genomic nucleotides were retained, a threshold demonstrated to yield high specificity.^41^ The list of all known humans pathogens was curated from taxonomic IDs compiled from the CZID compiled list^75^, the Bacterial and Viral Bioinformatics Resource Center (BV-BRC) list^76^ the Virulence Factor DataBase list^77^ and the Pathogen-Host Interactions database list.^78^ To confirm bacterial detection, we performed metagenomic assembly with Atlas under default parameters, with confirmation of contig identity by BLAST.^79^

### ARG abundance and diversity

For ARG abundance and diversity estimation, we used ARGprofiler (v1.0.0) with default settings.^52^ Briefly, ARGprofiler performs read quality control and trimming using *fastp* (v1.3.3), with the trimmed reads subsequently globally aligned with KMA(v1.4.12a)^80^ against the PanRes v1.0.2 reference database, which combines several existing databases (ResFinder, ResFinderFG, CARD, MegaRes, AMRFinderPlus and ARGANNOT) into one that includes more than 14000 unique ARGs.^52^ KMA reports the count of read fragments aligned to reference sequences. To control for mismapping, we filtered all results so that template coverage and identity was above 70%, with query identity and coverage above 90 and 70% respectively.

To infer the likely bacterial host and genomic context of the detected ARGs, we classified the KMA consensus sequence of each aligned reference gene using the CARD k-mer classifier implemented in RGI (v6.0.5) against CARD (v4.0.1).^81^ This approach matches query sequences against a set of 61-mers derived from CARD’s Resistomes and Variants dataset, in which AMR alleles are labelled by the taxa and genomic locations (chromosome or plasmid). Contigs were classified in FASTA mode (k = 61). As inputs were ARG contigs retained after the coverage and identity filtering described above, they were classified directly. A taxonomic and genomic assignment was made for each sequence exceeding the threshold of ten matching AMR k-mers, with the predicted host reported at species or genus level and genomic context classified as chromosomal, plasmid or either.

### Diversity analyses

Taxonomic diversity was calculated from datasets filtered by breadth-of-coverage (ESV) and relaxed unique k-mer thresholds (KU) as described above. For each canal and sampling date, reads were summed within taxon and profiles were computed at family and species-level resolution. Alpha diversity was estimated as the Shannon index on read-count profiles, with species richness taken as the number of taxa detected and Pielou’s evenness as the Shannon index divided by log richness. Differences in Shannon diversity across canals and across sampling dates were tested by Kruskal-Wallis tests, followed by pairwise Wilcoxon rank-sum tests with Benjamini-Hochberg correction. As viral species richness is sensitive to sequencing depth, we tested the association between richness and total read depth (Pearson, Spearman) and assessed whether canal-level richness differences persisted after linear adjustment for depth. Bray-Curtis dissimilarity was calculated between family-level relative-abundance (reads per million) profiles for each canal pair using scipy.spatial.distance separately for each sampling date.

For temporal trend analyses, species-level RPKMF values (that passed detection thresholds) were summed within the viral family per sample on the linear scale and log10-transformed. Family-level abundances were compared across timepoints using Kruskal-Wallis tests and, as all six sampling sites were sampled at every timepoint, canal-blocked Friedman and Page trend tests on per-canal mean abundances. Changes in detection frequency were assessed separately by exact permutation tests (20000 permutations) on the three by two timepoint-by-detection table. Families detected in fewer than five samples were excluded, and p-values were corrected across families using the Benjamini-Hochberg procedure. Families for which the minimum attainable permutation p-value exceeded the significance threshold (i.e. those with too few detections for any configuration of the data to yield a significant result) were excluded prior to multiple-testing correction. Sequencing depth and per-sample richness were tested for association with sampling week by Spearman correlation to exclude effort-driven artefacts.

### Estimating distance-decay relationships

Distance-decay effects were assessed by comparing physical distance in km to compositional similarity. Bray-Curtis dissimilarity was calculated between family-level relative abundance (RPM) profiles for each canal pair at each sampling date, and compositional similarity (was regressed against along-flow distance using Mantel tests (10000 label permutations constrained to preserve network connectivity) and Spearman correlations, on connected pairs only as well as on all pairs. The canal network forms a confluence in which two independent arms - a left arm (AJB>PET) and a right arm (AJA) - merge into a shared trunk (MB>EXP>CW). The pairs AJB-AJA and PET-AJA share no directed flow path. These were classified as hydrologically disconnected and the remaining 13 pairs as connected. As sequencing depth varied substantially between samples, and the terminal site CW had fewer than 1000 viral reads on two of three dates, all distance-decay analyses were repeated excluding these two samples as a depth-based sensitivity analysis.

### Estimating nestedness

For each sampling date, dissimilarity between canals was partitioned into turnover and nestedness-resultant components following the Baselga framework of Sørensen indices, at both family and species-level resolution and for both classifiers.^82^ Where multiple grab samples were collected within a canal, reads were pooled per canal before analysis. Containment was calculated for each directed upstream-downstream canal pair as the proportion of the upstream canal’s taxa also detected downstream, with a value of 1 indicating complete subsetting. Matrix-level nestedness was quantified using NODF (Nestedness metric based on Overlap and Decreasing Fill) and tested against a null distribution generated by the curveball algorithm (999 permutations), which preserves both richness and taxon prevalence. The relationship between richness and flow position was assessed by Spearman correlation. Containment and richness position analyses were restricted to hydrologically connected pairs. The Sorensen partition and NODF were computed across the full site-by-taxon matrix. For species-level analyses, NODF p-values were corrected for multiple comparisons across dates using Benjamini-Hochberg procedure. As presence/absence-based metrics are sensitive to sequencing effort, canals yielding fewer than 1000 viral reads on a date were excluded from that date’s analysis. The remaining canals were rarefied without replacement to the depth of the shallowest retained canal (100 independent draws), with a taxon considered present if detected in at least half of draws.

### Geographic shape files

Spatial data on the distribution of healthcare facilities were obtained from the GRID3 NGA - Health Facilities v2.0 dataset (https://data.grid3.org/). Catchment delineations and draining lines were obtained from es.world. Spatial data for mapping obtained from OpenStreetMap data, accessed through contextily library.

## Data availability

Sequence data is available on the European Nucleotide Archive (ENA) under project PRJEB120882.

## Code availability

All code to run the analyses is available at https://github.com/EdythParker/WW_lagos.

## Acknowledgements

The authors are grateful to the Lagos State Government and the Lagos State Waste Management office for their support in this project. The authors are also grateful for the support of the John D. and Catherine T. MacArthur Foundation, Flu Lab, and a cohort of generous donors through TED’s Audacious Project, including the ELMA Foundation, MacKenzie Scott, the Skoll Foundation, and Open Philanthropy. This work was also funded by NIH NIAID U01AI151812 (J.I.L).

## Author contributions

E.P., F.O., J.I.L., C.H. conceptualized the study. E.P., J.I.L., K.A., C.H. contributed the methodology. E.P., F.O., O.O., J.I.L. conducted the investigation. E.P., F.O. performed formal analysis. A.H., K,A. C.H. provided resources. E.P., F.O., H.S. curated data. E.P. wrote the original draft of the manuscript. All authors reviewed and edited the manuscript. E.P. performed visualization. J.I.L., C.H. supervised the study. F.O., J.I.L., undertook project administration. K.A., C.H. acquired funding.

