## Supplementary for "Wastewater metagenomic sequencing enables broad pathogen and resistome monitoring in Lagos, Nigeria"

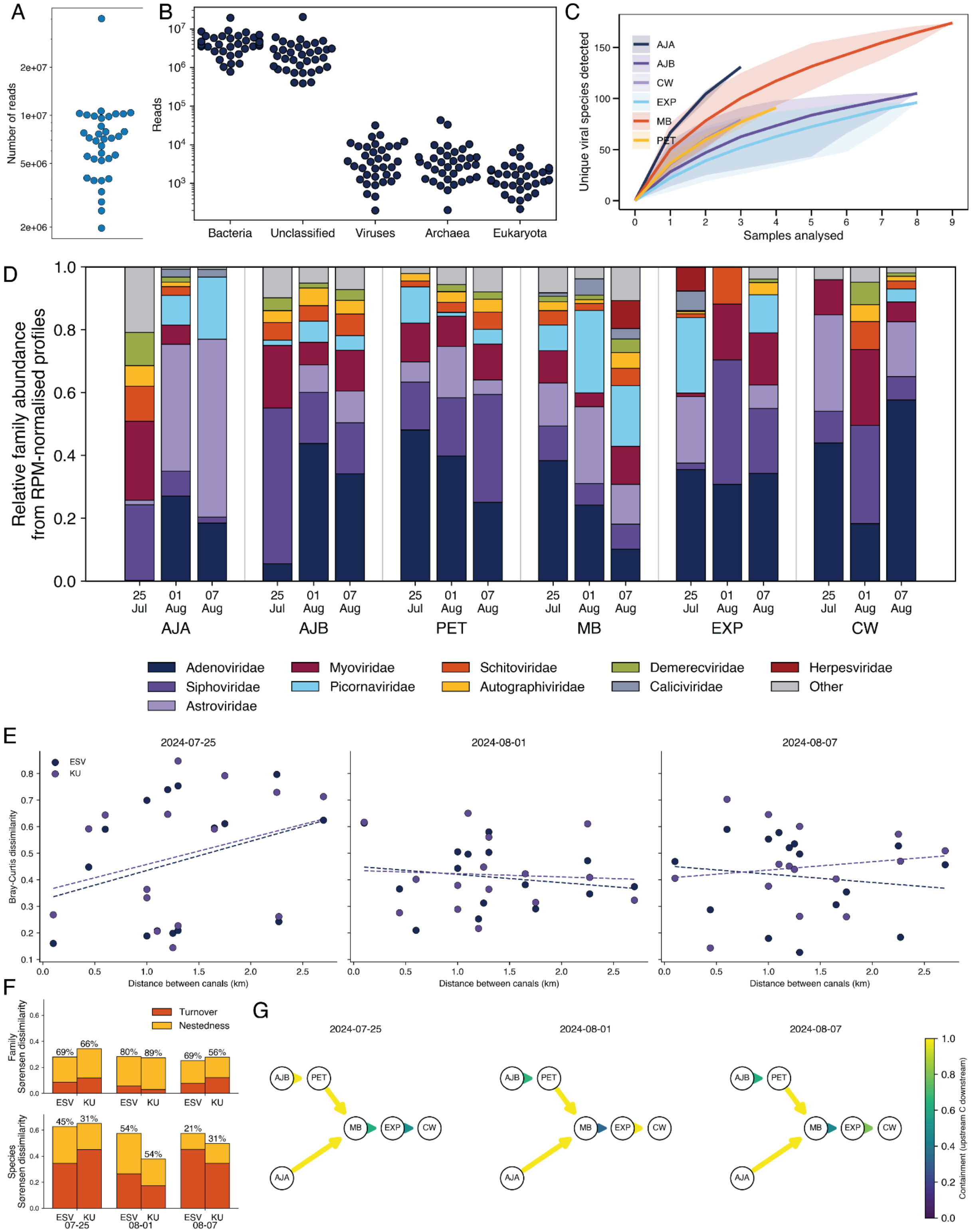


**Supplementary figure 1.** A) Distribution of total read counts across samples. B) Read assignments by domain, as per KU. C) Refraction curves for unique viral species identified across samples. Canal sampling points: Ajibade Babatola (AJB), Petrocam Canoe (PET), Ajao Estate (AJA), Mass Burial (MB), an Extra Point (EXP) and Canal Water (CW). D) Comparison of relative abundance of viral families across all three time points for each sampling site, as assigned by KU. E) Relationship between Bray-Curtis dissimilarity and physical distance for each canal-pair across time points. Method (Esviritu = ESV, KrakenUniq =KU) as per legend. F) Components of Sørensen dissimilarity at each time point across methods (ESV, KU) and taxonomic resolution. G) Containment flow diagram for species-level taxonomic resolution. The confluence is drawn as nodes (AJB, PET, AJA, MB, EXP, CW) connected by directed arrows along flow paths, with each arrow coloured/weighted by its containment value.

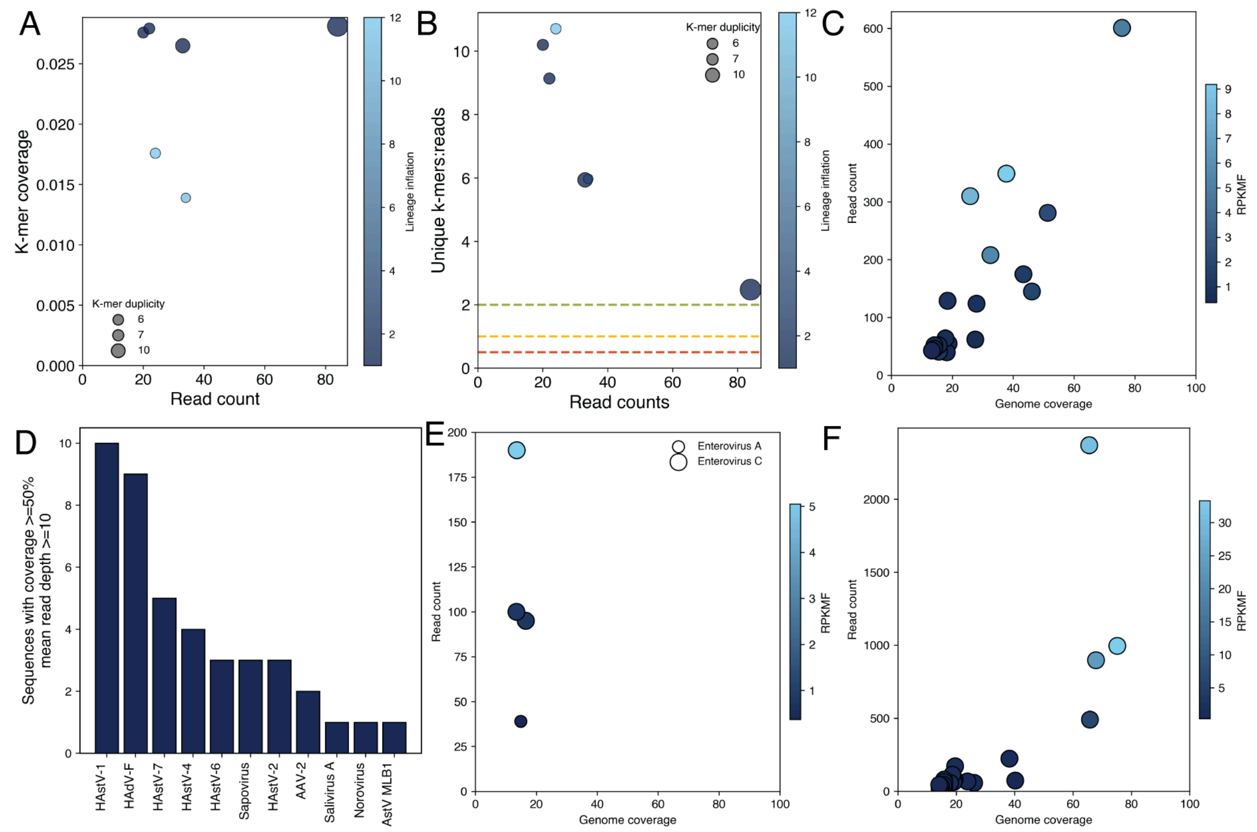


**Supplementary figure 2.** A) Relationship between K-mer coverage (coverage of the database k-mer space) and assigned read counts for Noroviruses, as per KU. Points are coloured by lineage inflation, defined as the ratio of total clade-level reads to reads assigned directly to the species node, and sized by k-mer duplicity, the multiplicity with which the same k-mers are sampled (or duplicated). B) The relationship between the ratio of unique k-mers to reads counts and the number of reads for Noroviruses, as per KU. Points are coloured by lineage inflation, and sized by k-mer duplicity. C) The relationship between read count and genome breadth of coverage for Noroviruses. Points are coloured by RPKMF, as per ESV. D) The number of contigs with genome coverage above 50% and mean read depth above 10. E) The relationship between read count and genome breadth of coverage for Enteroviruses, as per legend. Points are coloured by RPKMF, as per ESV. F) The relationship between read count and genome breadth of coverage for Sapoviruses. Points are coloured by RPKMF, as per ESV.
